# Social distancing strategies for curbing the COVID-19 epidemic

**DOI:** 10.1101/2020.03.22.20041079

**Authors:** Stephen Kissler, Christine Tedijanto, Marc Lipsitch, Yonatan H. Grad

## Abstract

The SARS-CoV-2 pandemic is straining healthcare resources worldwide, prompting social distancing measures to reduce transmission intensity. The amount of social distancing needed to curb the SARS-CoV-2 epidemic in the context of seasonally varying transmission remains unclear. Using a mathematical model, we assessed that one-time interventions will be insufficient to maintain COVID-19 prevalence within the critical care capacity of the United States. Seasonal variation in transmission will facilitate epidemic control during the summer months but could lead to an intense resurgence in the autumn. Intermittent distancing measures can maintain control of the epidemic, but without other interventions, these measures may be necessary into 2022. Increasing critical care capacity could reduce the duration of the SARS-CoV-2 epidemic while ensuring that critically ill patients receive appropriate care.

**Summary:** One-time distancing results in a fall COVID-19 peak. Intermittent efforts require greater hospital capacity and surveillance.

## Full text

The COVID-19 pandemic is causing substantial mortality and a major strain on healthcare systems *(1,2)*. With no pharmaceutical treatments available, interventions have focused on contact tracing, quarantine, and social distancing. Intensive testing, tracing, and isolation of cases has enabled control of transmission in some places, such as Singapore and Hong Kong *(3)*. At the opposite extreme, many countries lack the testing and public health resources to mount similar responses to the COVID-19 epidemic, which could result in unhindered spread and catastrophic outbreaks. Between these responses, many countries are adopting measures termed “social distancing” or “physical distancing,” closing schools and workplaces and limiting the sizes of gatherings. The goal of these strategies is to slow the spread of infection and reduce the intensity of the epidemic (“flatten the curve”) *(3)*, thus reducing risk of overwhelming health systems and buying time to develop treatments and vaccines. However, there is concern that such measures may have to stay in place over long periods, perhaps with occasional periods of loosening, as immunity slowly accumulates in the population to a point where it would preclude future outbreaks.

The intensity (peak size) of outbreaks depends strongly on the degree of seasonal forcing of transmission *(4)*. The transmission of many respiratory pathogens, including the human coronaviruses that cause mild common cold-like syndromes, is seasonal in temperate regions, peaking in the winter months *(4)*. This variation in transmission strength may be driven by a variety of factors, including increased indoor crowding in the winter, the onset of the school term in the autumn, and climate factors *(5)*. If SARS-Cov-2 transmission is similarly subject to seasonal forcing, summer outbreaks would naturally have lower peaks than winter outbreaks. This aligns with observations from influenza pandemics, where relatively small spring and summer outbreaks are frequently followed by larger autumn/winter outbreaks *(6)*. Due to seasonal variation in transmission strength, it may be more difficult to flatten epidemic curves in the winter than in the summer. Moreover, a winter peak for COVID-19 will coincide with peak influenza *(5)*, further straining health care systems.

A key metric for the success of curve-flattening strategies is whether critical care capacities are exceeded. Modeling studies *(7)* and experience from the Wuhan outbreak *(2)* indicate that critical care capacities even in high-income countries can be exceeded many times over if distancing measures are not implemented quickly or strongly enough. To keep critical care capacities from being overwhelmed, prolonged or intermittent social distancing may be necessary *(7)*. However, the necessary duration, frequency, and intensity of this distancing remains unclear in the presence of seasonality. To alleviate these problems, approaches to increase critical care capacity have included rapid construction or repurposing of hospital facilities and consideration of increased manufacturing and distribution of ventilators *(8–11)*.

The duration of social distancing measures needed to maintain control of the SARS-CoV-2 epidemic in the context of seasonally varying transmission remains unclear, as does the impact of increasing critical care capacity on the overall trajectory of the outbreak. To address these unknowns, here we assessed the transmission of SARS-Cov-2 through 2021 using a mathematical model with seasonal transmission forcing.

We used a deterministic (ordinary differential equation) mathematical model to simulate the transmission of SARS-CoV-2. The model is an adapted Susceptible-Exposed-Infectious-Recovered (SEIR) model with three tracks (**Figure S1)** to account for individuals who are asymptomatic or have mild symptoms (95.6%), individuals who are hospitalized but do not require critical care (3.08%), and individuals who are hospitalized and require critical care (1.32%) *(7)*. The mean incubation period was 4.6 days, the infectious period/mean time to hospitalization was 4 days, the mean duration of non-critical hospital stay was 8 days for those not requiring critical care and 6 days for those requiring critical care, and the mean duration of critical care was 10 days *(7)*. Seasonal forcing was incorporated by allowing the basic reproduction number (*R*_*0*_) to follow a cosine curve that peaks in early December *(4)*. We varied the peak (wintertime) *R*_*0*_ between 2 and 2.5 and allowed the summertime *R*_*0*_ to vary between 70% and 100% (*i*.*e*. no seasonality) of the wintertime *R*_*0*_ *(4)*.

We used the open critical care capacity of the United States, 0.89 free beds per 10,000 adults, as a benchmark for critical care demand *(2)*. We simulated epidemic trajectories based on an epidemic establishment time of 11 March 2020. We simulated social distancing by reducing *R*_*0*_ by a fixed proportion, which ranged between 0 and 60%, on par with the reduction in *R*_*0*_ achieved in China through intense social distancing measures *(3)*. We assessed ‘one-time’ social distancing interventions, for which *R*_*0*_ was reduced by up to 60% for a fixed duration of time (up to 20 weeks) starting two weeks after epidemic establishment. We also assessed intermittent social distancing measures, for which social distancing was turned ‘on’ when the prevalence of infection rose above a threshold and ‘off’ when it fell below a second, lower threshold, with the goal of keeping the number of critical care patients below 0.89 per 10,000 adults. An ‘on’ threshold of 37.5 cases per 10,000 people achieved this goal in both the seasonal and non-seasonal cases with wintertime *R*_*0*_ = 2. We chose 10 cases per 10,000 adults as the ‘off’ threshold. We performed a sensitivity analysis around these threshold values (**Figures S2-S3**) to assess how they affected the duration and frequency of the interventions. Finally, we assessed the impact of doubling critical care capacity (and the associated on/off thresholds) on the frequency and overall duration of the period social distancing measures.

We evaluated the impact of one-time social distancing efforts of varying effectiveness and duration on the peak and timing of the epidemic with and without seasonal forcing. When transmission is not subject to seasonal forcing, one-time social distancing measures reduce the epidemic peak size (**Figure 1, Figure S4**). Under all scenarios, there was a resurgence of infection when the simulated social distancing measures were lifted. However, longer and more stringent social distancing did not always correlate with greater reductions in epidemic peak size. In the case of a 20-week period of social distancing with 60% reduction in *R*_*0*_, for example, the resurgence peak size is nearly the same as the peak size of the uncontrolled epidemic: the social distancing is so effective that virtually no population immunity is built. The greatest reductions in peak size come from social distancing intensity and duration that divide cases approximately equally between peaks *(12)*.

**Figure 1.**
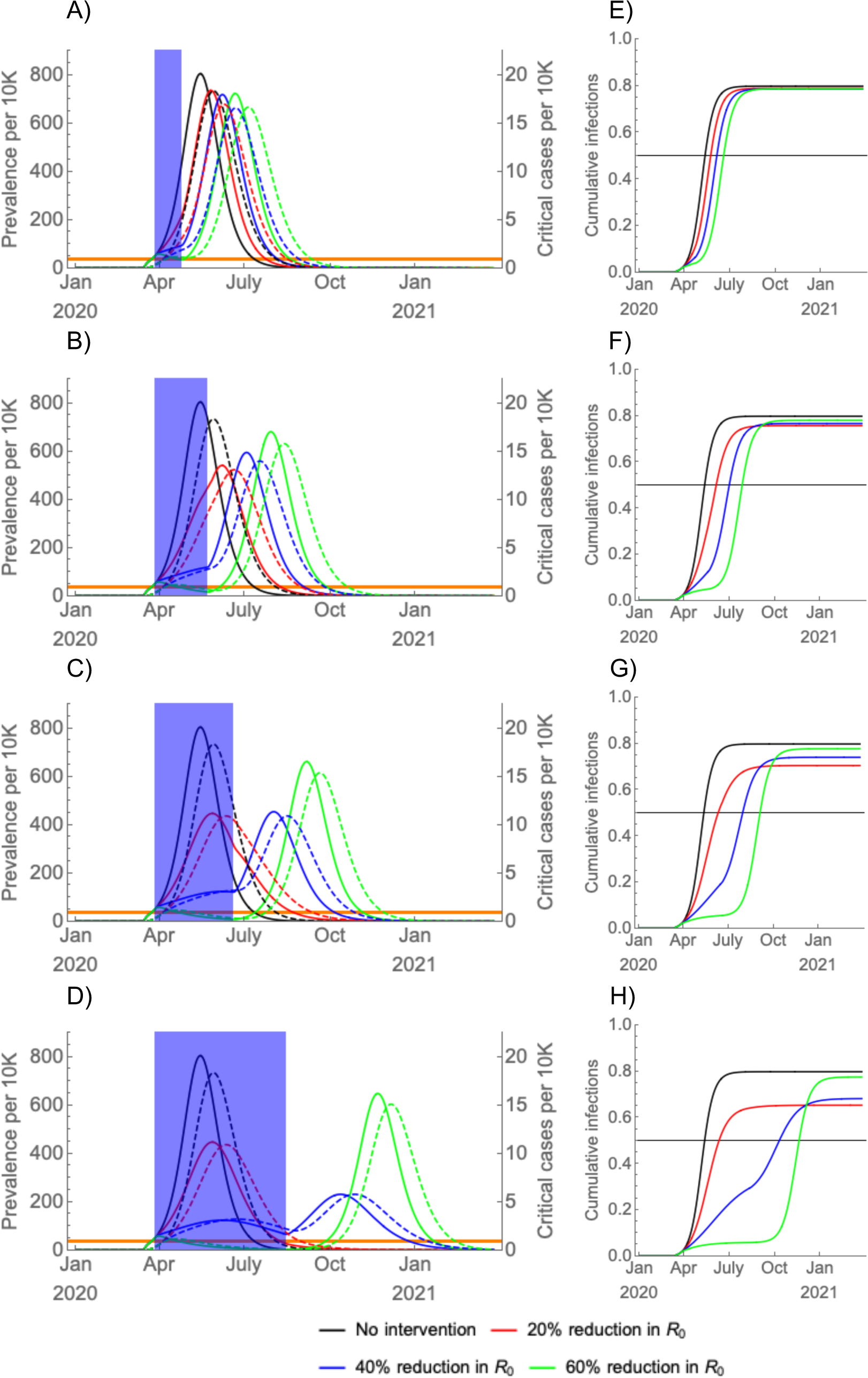
(A-D) Simulated prevalence of COVID-19 infections (solid) and critical COVID-19 cases (dashed) following establishment on 11 March 2020 with a period of social distancing (shaded blue region) instated two weeks later, with the duration of social distancing lasting (A) four weeks, (B) eight weeks, (C) twelve weeks, and (D) twenty weeks. There is no seasonal forcing; R_0_ was held constant at 2 (see **Figure S4** for *R*_*0*_ = 2.5). The effectiveness of social distancing varied from none to a 60% reduction in *R*_*0*_. Cumulative infection sizes are depicted beside each prevalence plot (E-H) with the herd immunity threshold (horizontal black bar). Long-term (20-week), moderately effective (20%-40%) social distancing yields the smallest overall peak and total outbreak size.

For simulations with seasonal forcing, the post-intervention resurgent peak could exceed the size of the unconstrained epidemic (**Figure 2, Figure S5**), both in terms of peak prevalence and in terms of total number infected. Strong social distancing maintains a high proportion of susceptible individuals in the population, leading to an intense epidemic when *R*_*0*_ rises in the late autumn and winter. None of the one-time interventions was effective in maintaining the prevalence of critical cases below the critical care capacity.

**Figure 2.**
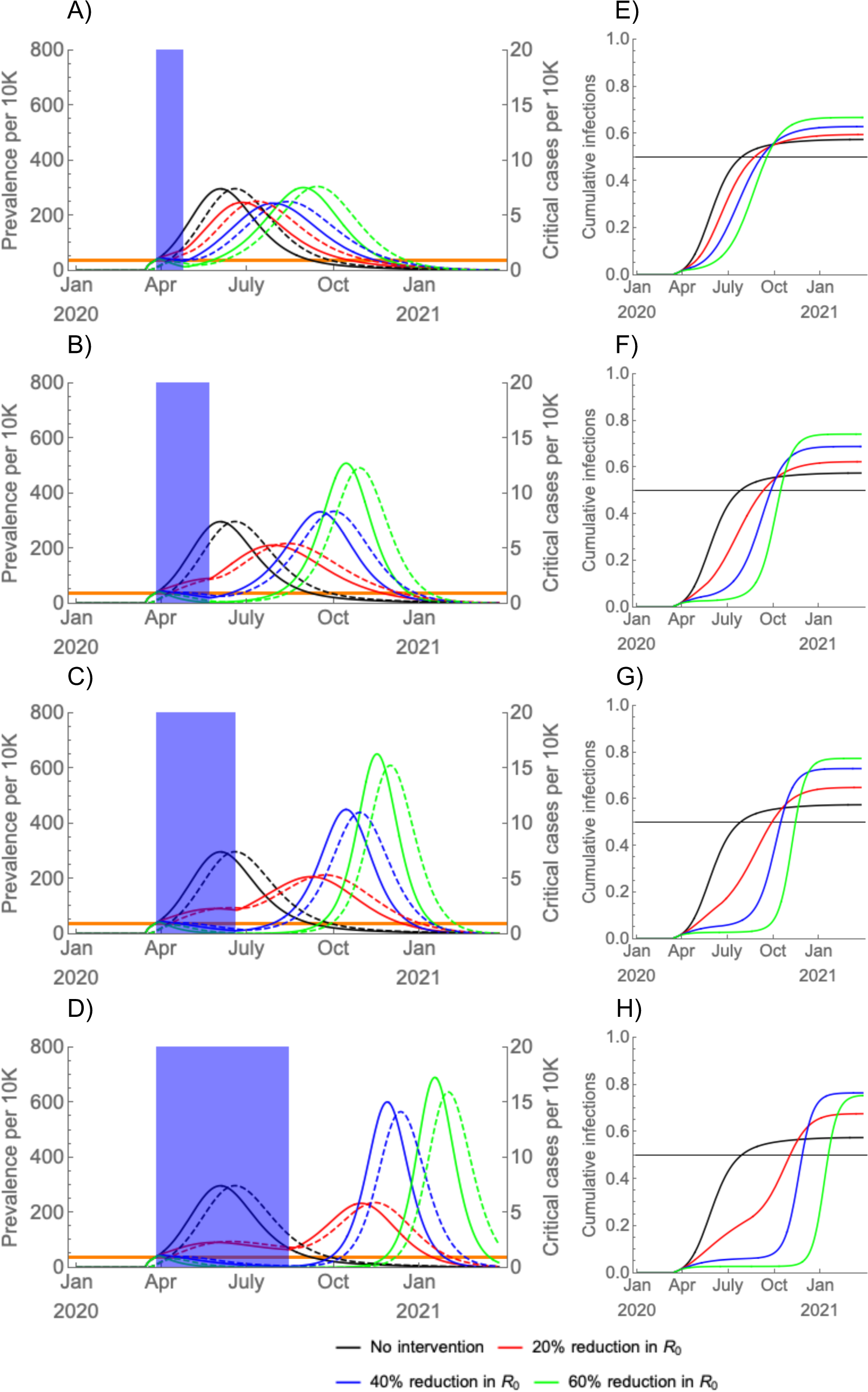
(A-D) Simulated prevalence, assuming seasonal forcing (wintertime *R*_*0*_ = 2, summertime *R*_*0*_ = 1.4), of COVID-19 infections (solid) and critical COVID-19 cases (dashed) following establishment on 11 March 2020 with a period of social distancing (shaded blue region) instated two weeks later, with the duration of social distancing lasting (A) four weeks, (B) eight weeks, (C) twelve weeks, and (D) twenty weeks (see **Figure S5** for a scenario with wintertime *R*_*0*_ = 2.5). The effectiveness of social distancing varied from none to a 60% reduction in *R*_*0*_. Cumulative infection sizes are depicted beside each prevalence plot (E-H) with the herd immunity threshold (horizontal black bar). Preventing widespread infection during the summer can flatten and prolong the epidemic but can also lead to a high density of susceptible individuals who could become infected in an intense autumn wave.

Intermittent social distancing can prevent critical care capacity from being exceeded (**Figure 3, Figure S6**). Due to the natural history of infection, there is an approximately 3-week lag between the start of social distancing and the peak critical care demand. When transmission is seasonally forced, summertime social distancing can be less frequent than when *R*_*0*_ remains constant at its maximal wintertime value throughout the year. The length of time between distancing measures increases as the epidemic continues, as the accumulation of immunity in the population slows the resurgence of infection. Under current critical care capacities, however, the overall duration of the SARS-CoV-2 epidemic could last into 2022, requiring social distancing measures to be in place between 25% (for wintertime *R*_*0*_ = 2 and seasonality, **Figure S3A**) and 70% (for wintertime *R*_*0*_ = 2.5 and no seasonality, **Figure S2C**) of that time.

**Figure 3.**
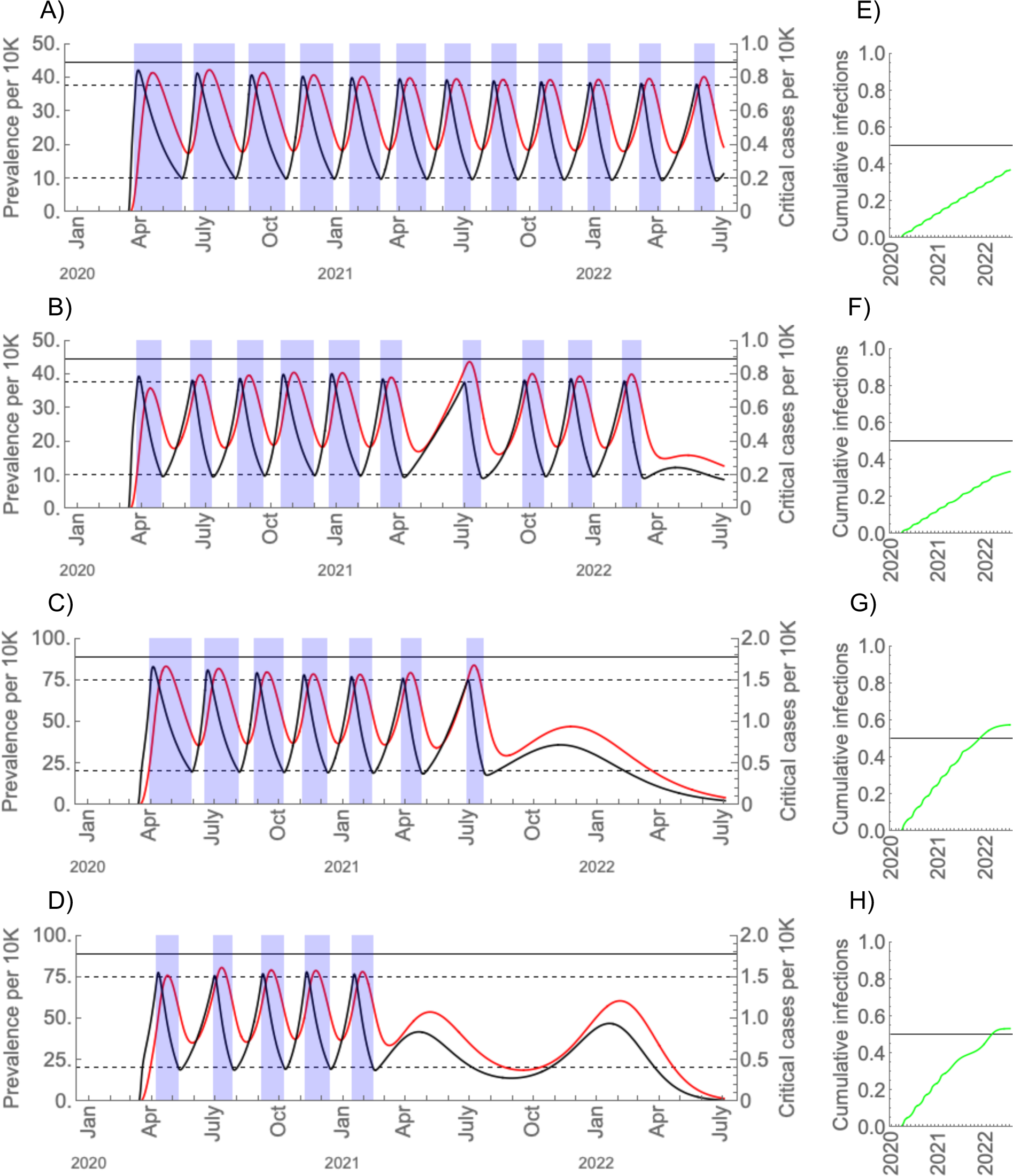
SARS-Cov-2 prevalence (black curves) and critical cases (red curves) under intermittent social distancing (shaded blue regions) without seasonal forcing (A, C) and with seasonal forcing (B, D). Distancing yields a 60% reduction in *R*_*0*_. Critical care capacity is depicted by the solid horizontal black bars; (A) and (B) are the scenarios with current US critical care capacity and (C) and (D) are the scenarios with double the current critical care capacity. The maximal wintertime *R*_*0*_ is 2 and for the seasonal scenarios the summertime *R*_*0*_ is 1.4. Prevalence is in black and critical care cases are in red. To the right of each main plot (E-H), the proportion immune over time is depicted in green with the herd immunity threshold (horizontal black bar).

Increasing critical care capacity allows population immunity to be accumulated more rapidly, reducing the overall duration of the epidemic and the total length of social distancing measures (**Figure 3 C-D**). While the frequency and duration of the social distancing measures is similar between the scenarios with current and expanded critical care capacity, the epidemic concludes by July 2022 and social distancing measures can be fully relaxed by early- to mid-2021, depending again on the degree of seasonal forcing of transmission (**Figure 3 C-D**). We anticipate that SARS-CoV-2 will then circulate seasonally with winter peaks in subsequent years *(4)*.

A single period of social distancing will not be sufficient to prevent critical care capacities from being overwhelmed by the COVID-19 epidemic, because under any scenario considered it leaves enough of the population susceptible that a rebound in transmission after the end of the period will lead to an epidemic that exceeds this capacity. This resurgence could be especially intense if it coincides with a wintertime rise in *R*_*0*_. Intermittent social distancing can maintain the prevalence of critical COVID-19 illness within current capacities, but this strategy could prolong the overall duration of the epidemic into 2022. Increasing critical care capacities would substantially reduce the overall duration of the epidemic while ensuring adequate care for the critically ill.

Our findings agree with observational and modelling studies *(2,7)* that find that early implementation of strong social distancing is essential for controlling the spread of SARS-CoV-2 and that, in the absence of the development of new therapies or preventative measures, such as aggressive case finding and quarantining *(13)*, intermittent distancing measures may be the only way to avoid overwhelming critical care capacity while building population immunity. The observation that strong social distancing can lead to especially large resurgences agrees with data from the 1918 influenza pandemic in the United States *(14)*, in which the size of the autumn 1918 peak of infection was inversely associated with that of a subsequent winter peak after interventions were no longer in place.

To implement an effective intermittent social distancing strategy, it will be necessary to carry out widespread surveillance to monitor when the prevalence thresholds that trigger the beginning or end of distancing have been crossed. Without such surveillance, critical care bed availability might be used as a proxy for prevalence, but this metric is far from optimal since the lag between distancing and peak critical care demand could lead to frequent overrunning of critical care resources. Under some circumstances, intense social distancing may be able to reduce the prevalence of COVID-19 enough to warrant a shift in strategy to contact tracing and containment efforts, as has occurred in many parts of China *(13,15,16)*. Still, countries that have achieved this level of control of the outbreak should prepare for the possibility of substantial resurgences of infection and a return to social distancing measures, especially if seasonal forcing contributes to a rise in transmissibility in the winter.

Treatments or vaccines for SARS-CoV-2 would reduce the duration and intensity of social distancing required to maintain control of the epidemic. Treatments could reduce the proportion of infections that require critical care and could reduce the duration of infectiousness, which would both directly and indirectly (through a reduction in *R*_*0*_) reduce the demand for critical care resources. A vaccine would accelerate the accumulation of immunity in the population, reducing the overall length of the epidemic and averting infections that might have resulted in a need for critical care. Still, the development and widespread adoption of pharmaceutical interventions will take months at best, so a period of sustained or intermittent social distancing will almost certainly be necessary.

The model we have used is deterministic and assumes that the population is well-mixed, meaning that each person has an equal probability of coming into potentially infectious contact with every other person. Population structure can reduce outbreak peak size *(17)*, so the modeling here may be interpreted as worst-case scenarios. The timing and duration of the interventions will likely need to be tailored geographically due to variation in epidemic timing, seasonal forcing, and critical care capacities. Deterministic models also cannot capture the possibility of SARS-CoV-2 elimination. However, given the widespread transmission of SARS-CoV-2, we believe that elimination is unlikely. Re-introductions of SARS-CoV-2 from locations with ongoing outbreaks would be sufficient to initiate sustained local transmission again *(18)*. We have also assumed that SARS-CoV-2 infection is permanently immunizing, since our focus was on the initial pandemic period. There is insufficient serological data to assess the amount of immunity that exists to SARS-CoV-2 and its duration. If SARS-CoV-2 immunity wanes rapidly, social distancing measures may need to be extended longer. However, if there are many undocumented asymptomatic infections that lead to immunity *(19)*, less social distancing may be required. Social distancing strategies will require further evaluation as longitudinal serological studies clarify the extent and duration of immunity to SARS-Cov-2.

One-time social distancing efforts will push the SARS-CoV-2 epidemic peak into the autumn, potentially exacerbating the load on critical care resources if there is increased wintertime transmissibility. Intermittent social distancing can maintain critical care demand within current thresholds, but widespread surveillance will be required to time the distancing measures correctly and avoid overshooting critical care capacity. New therapeutics, vaccines, or other interventions such as aggressive contact tracing and quarantine – impractical now in many places but more practical once case numbers have been reduced and testing scaled up *(20)* – could alleviate the need for stringent social distancing to maintain control of the epidemic. In the absence of such interventions, surveillance and intermittent distancing may need to be maintained into 2022, which would present a substantial social and economic burden. To shorten the SARS-CoV-2 epidemic and ensure adequate care for the critically ill, increasing critical care capacity and developing additional interventions are urgent priorities.

## Data Availability

The manuscript includes no data.

## Supplement

**Figure S1.**
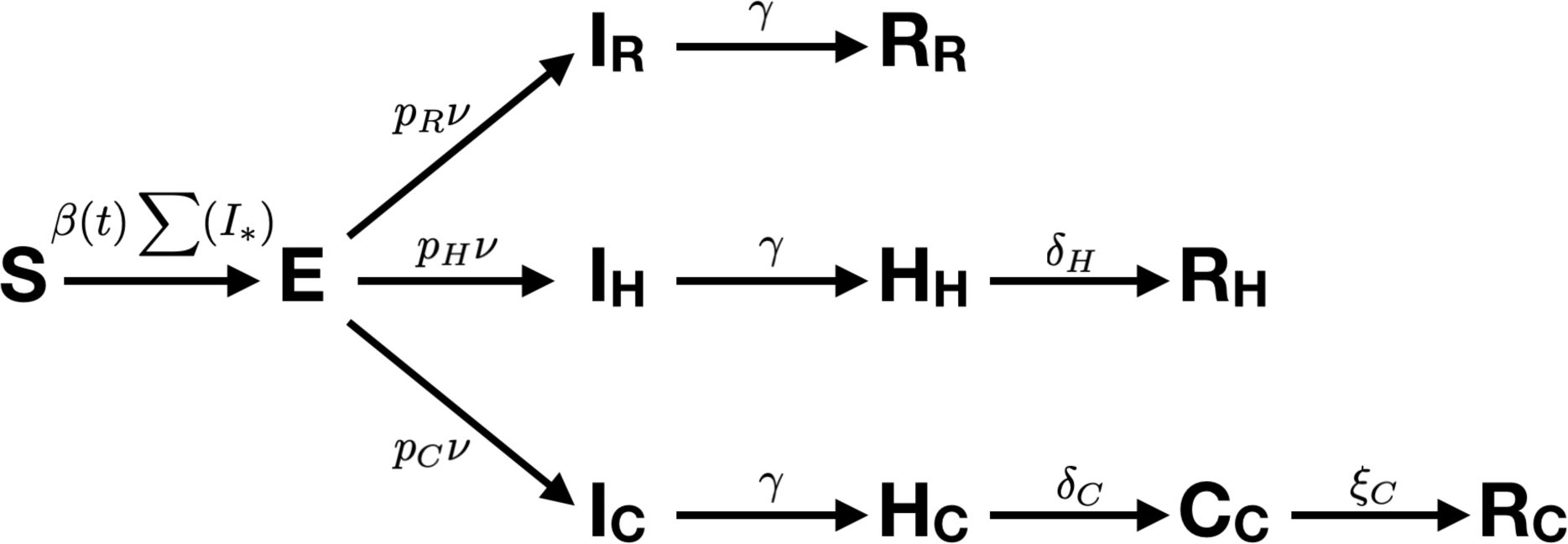
Schematic diagram of the disease transmission model. The population begins as susceptible (S). Infection is introduced through a half-week pulse in the force of infection starting on 11 Mar 2020*(4)*. The transmission rate *β(t)* is a cosine with 52-week period parametrized by a phase shift (*φ*), a maximum value (max(*R*_*0*_)), and a minimum value (*Δ\** max(*R*_*0*_)).Infected individuals then proceed to an exposed (E) state, after which a proportion *p*_*R*_ enters the ‘recovery’ arm, *p*_*H*_ enters the ‘hospitalization’ arm, and *p*_*C*_ enters the ‘critical care’ arm. Exposed individuals become infectious (*I*) at rate *ν*. Individuals in the recovery arm then recover *(R*), but individuals in the hospitalization arm must pass through the hospitalization state (*H*) and individuals in the critical care arm must pass through both the hospitalization (*H*) and critical care *(C*) states. Parameter values are given in Table S1.

**Figure S2.**
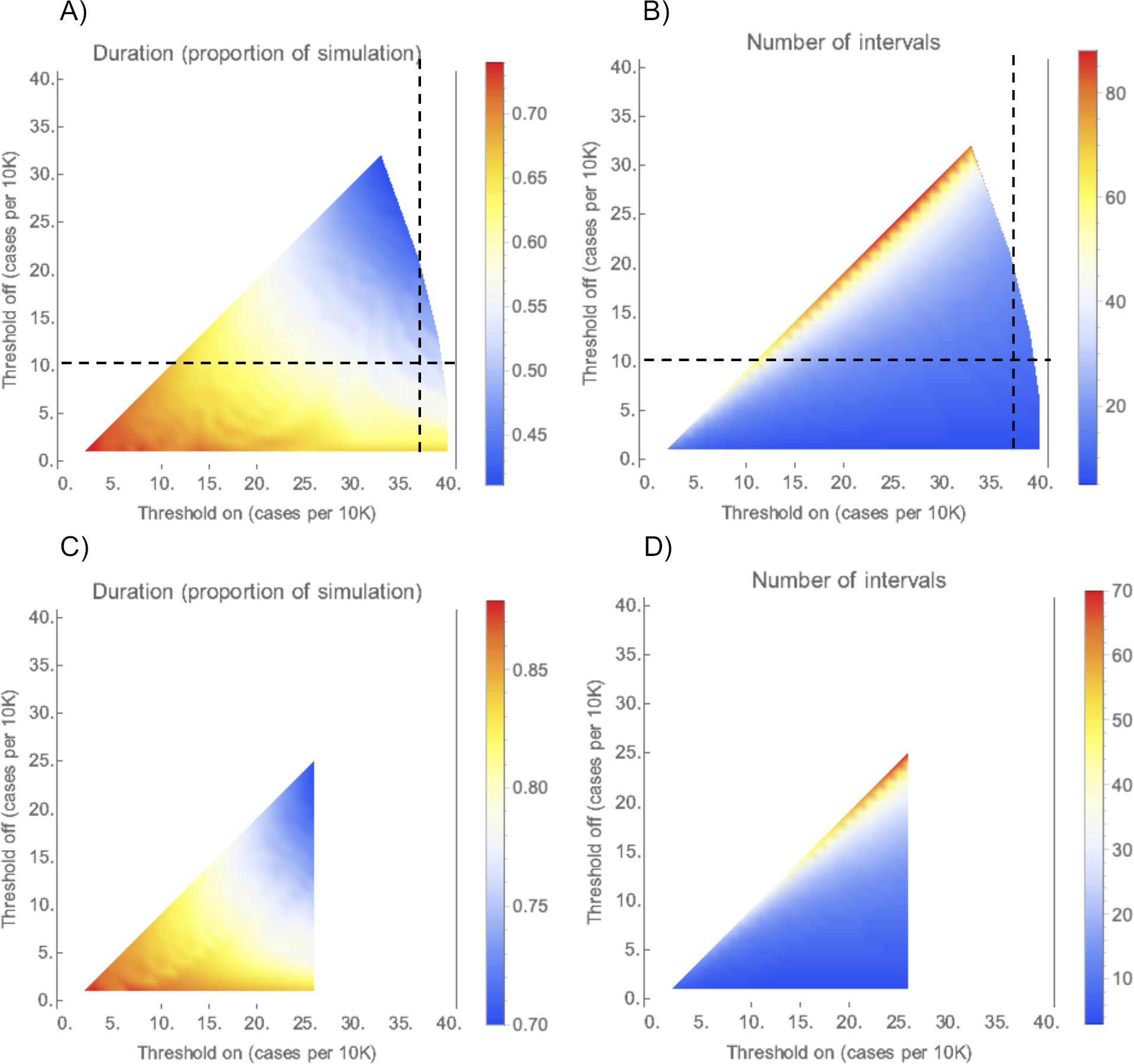
Duration (A, C) and number (B, D) of social distancing intervals for ‘on’ and ‘off’ thresholds between 1 and 40 cases per 10,000 people from simulations with no seasonal variation in transmission. Simulations were run from 1 Jan 2020 through 1 July 2022, with an epidemic establishment time on 11 March 2020. For sub-figures A and B, *R*_*0*_ is For sub-figures C and D, *R*_*0*_ is 2.5 (note the difference in color scales). The ‘on’ threshold must be greater than the ‘off’ threshold (leaving the top-left region of each plot blank) and the number of critical care cases cannot exceed the current US capacity of 0.89 per 10,000 people (leaving the right-hand region of each plot blank). The dashed lines in sub-figures A and B mark the threshold values used to produce **Figure 3A**.

**Figure S3.**
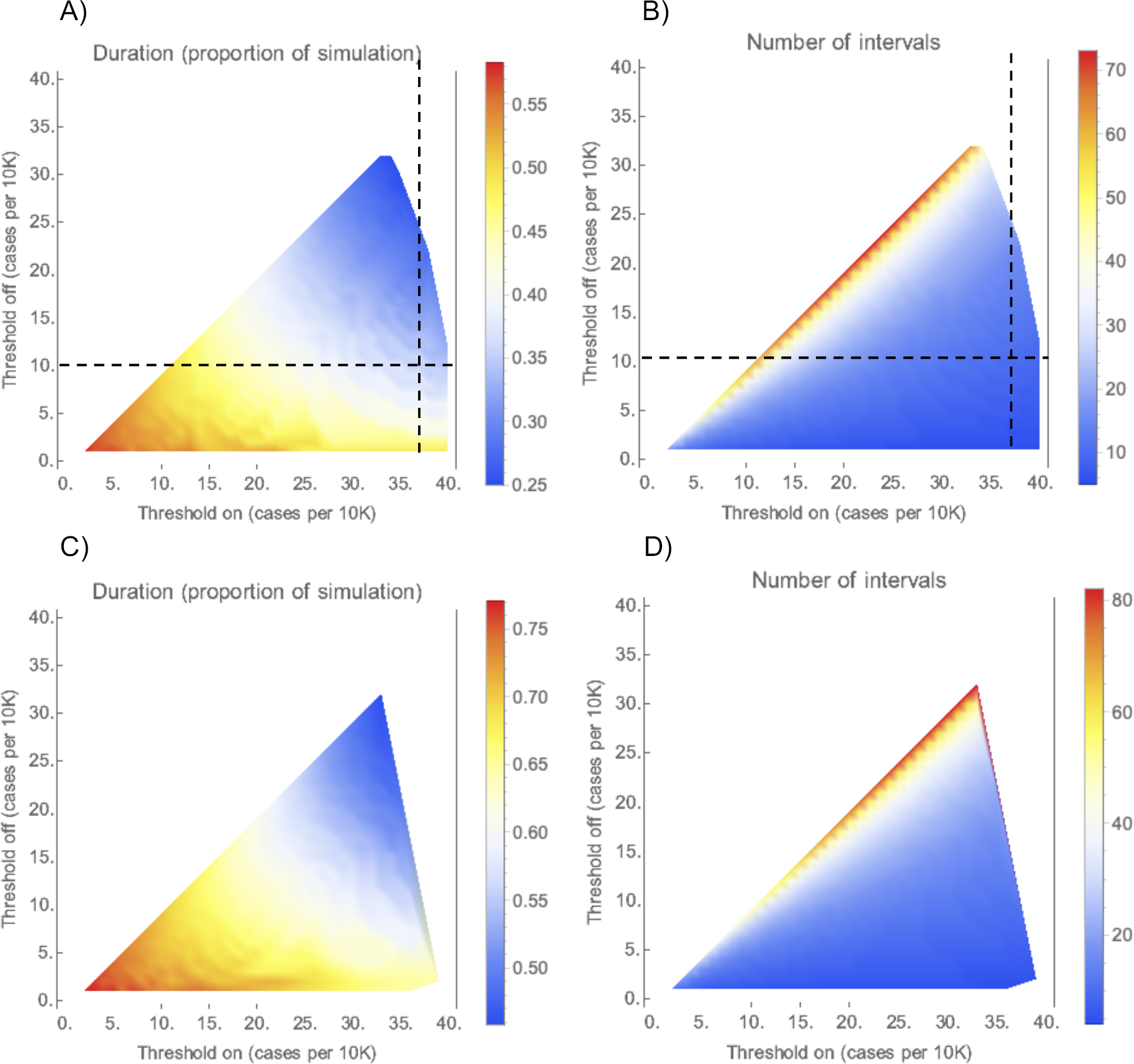
Duration (A, C) and number (B, D) of social distancing intervals for ‘on’ and ‘off’ thresholds between 1 and 40 cases per 10,000 people from simulations with seasonal transmission. Simulations were run from 1 Jan 2020 through 1 July 2022, with an epidemic establishment time on 11 March 2020. For sub-figures A and B, the wintertime *R*_*0*_ is 2 and the summertime *R*_*0*_ is 1.4. For sub-figures C and D, the wintertime *R*_*0*_ is 2.5 and the summertime *R*_*0*_ is 1.75 (note the difference in color scales). The ‘on’ threshold must be greater than the ‘off’ threshold (leaving the top-left region of each plot blank) and the number of critical care cases cannot exceed the current US capacity of 0.89 per 10,000 people (leaving the right-hand region of each plot blank). The dashed lines in sub-figures A and B mark the threshold values used to produce **Figure 3B**.

**Figure S4.**
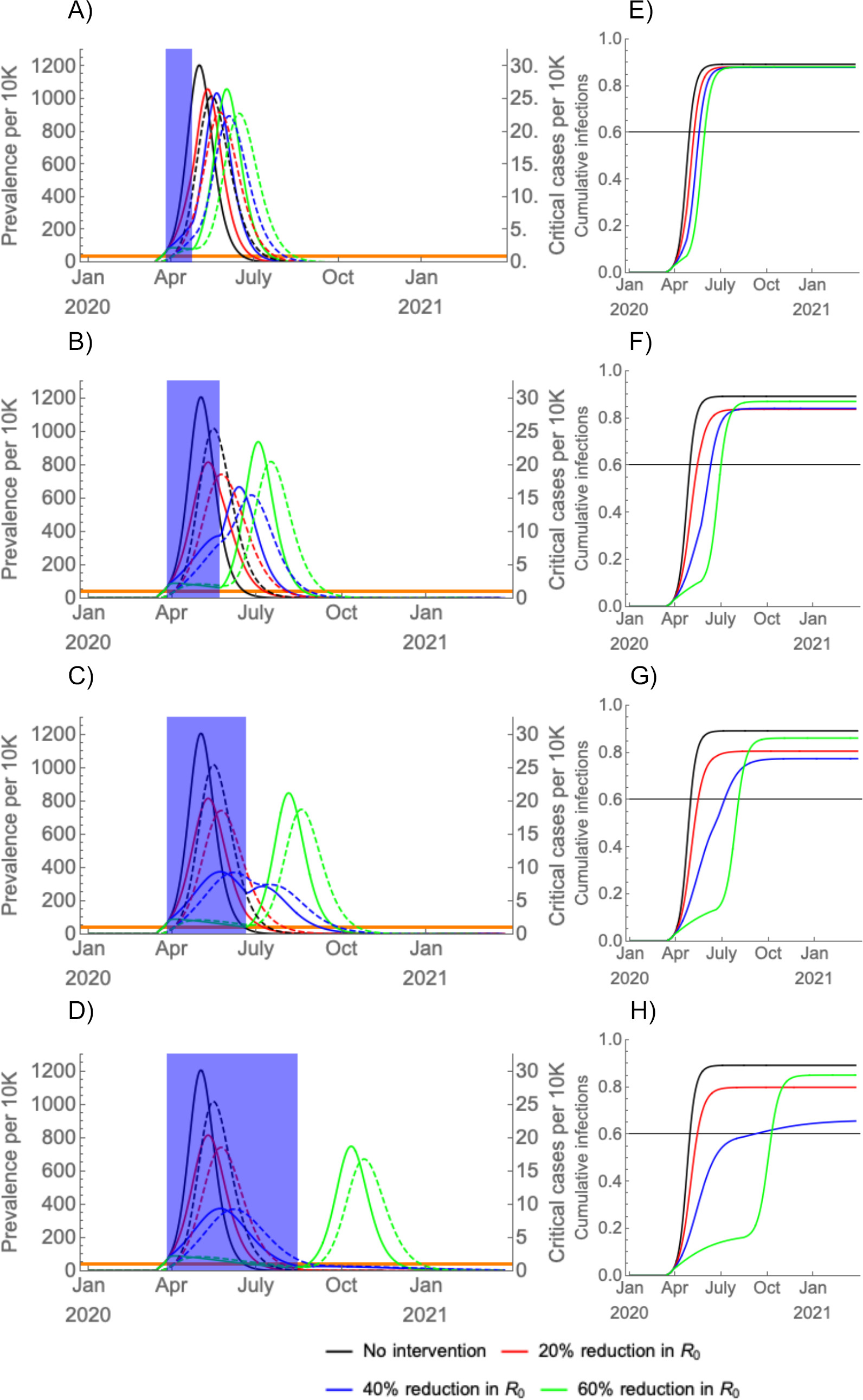
(A-D) Simulated prevalence of COVID-19 infections (solid) and critical COVID-19 cases (dashed) following establishment on 11 March 2020 with a period of social distancing (shaded blue region) instated two weeks later, with the duration of social distancing lasting (A) four weeks, (B) eight weeks, (C) twelve weeks, and (D) twenty weeks. There is no seasonal forcing; R_0_ was held constant at 2.5. The effectiveness of social distancing varied from none to a 60% reduction in *R*_*0*_. Cumulative infection sizes are depicted beside each prevalence plot (E-H) with the herd immunity threshold (horizontal black bar). Long-term (20-week), moderately effective (40%) social distancing yields the smallest overall peak and total outbreak size.

**Figure S5.**
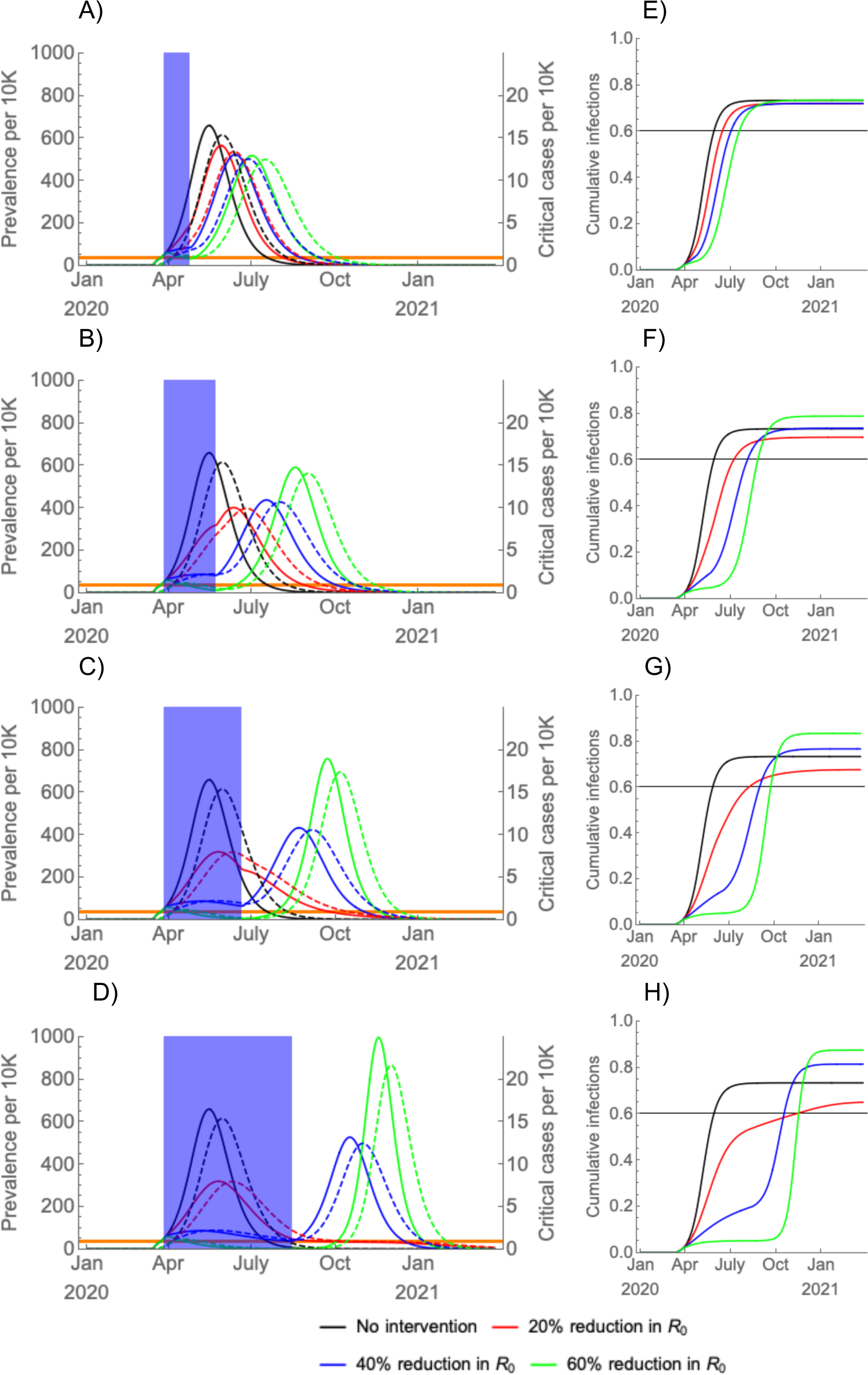
(A-D) Simulated prevalence, assuming seasonal forcing (wintertime *R*_*0*_ = 2.5, summertime *R*_*0*_ = 1.75), of COVID-19 infections (solid) and critical COVID-19 cases (dashed) following establishment on 11 March 2020 with a period of social distancing (shaded blue region) instated two weeks later, with the duration of social distancing lasting (A) four weeks, (B) eight weeks, (C) twelve weeks, and (D) twenty weeks. The effectiveness of social distancing varied from none to a 60% reduction in *R*_*0*_. Cumulative infection sizes are depicted beside each prevalence plot (E-H) with the herd immunity threshold (horizontal black bar). Preventing widespread infection during the summer can flatten and prolong the epidemic but can also lead to a high density of susceptible individuals who could become infected in an intense autumn wave.

**Figure S6.**
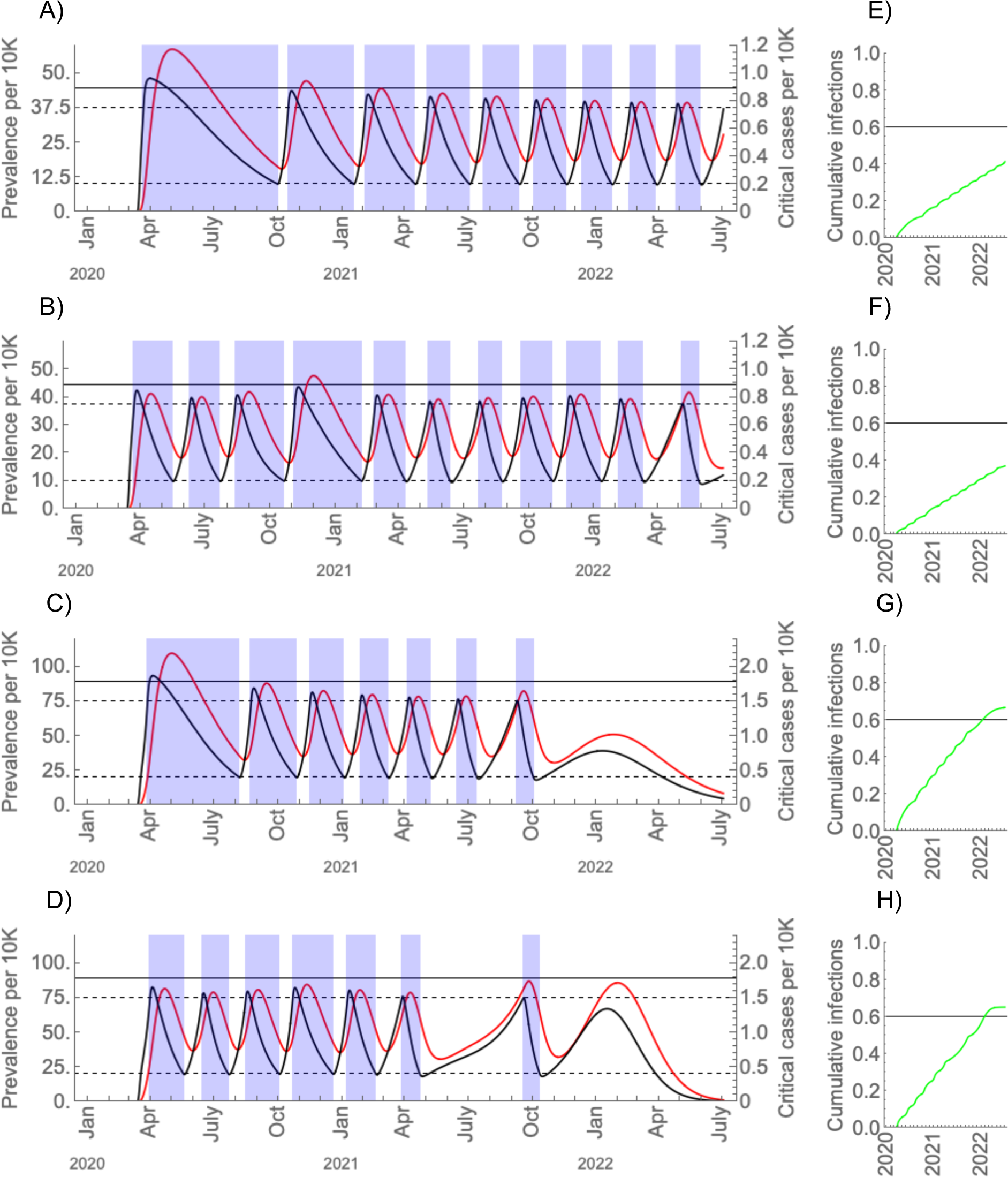
Intermittent social distancing (shaded blue regions) without seasonal forcing (A, C) and with seasonal forcing (B, D) with current critical care capacity (A, B; solid black bar) and double the current critical care capacity (C, D; solid black bar). The maximal wintertime *R*_*0*_ is 2.5 and for the seasonal scenarios the summertime *R*_*0*_ is 1.75. Distancing yields a 60% reduction in *R*_*0*_. Prevalence is in black and critical care cases are in red. To the right of each main plot (E-H), the proportion immune over time is depicted in green with the herd immunity threshold (horizontal black bar).

**Table S1.** Parameter values for the transmission model, from *(4,7)*.

| Parameter | Value/Range | Units | Meaning |
| --- | --- | --- | --- |
| $p_R$ | 0.956 | None | Proportion of infections that are not hospitalized |
| $p_H$ | 0.0308 | None | Proportion of infections that are hospitalized (excluding critical care) |
| $p_C$ | 0.0132 | None | Proportion of infections that receive critical care |
| $1/\gamma$ | 5 | Days | Infectious period/period prior to hospitalization |
| $1/\delta_H$ | 8 | Days | Duration of hospitalization for cases that do not receive critical care |
| $1/\delta_C$ | 6 | Days | Duration of hospitalization prior to critical care |
| $1/\xi_C$ | 10 | Days | Duration of critical care |
| $\max(R_0)$ | [2, 2.5] | None | Basic reproduction number |
| $\Delta$ | [0.7, 1.0] | None | Proportional decline in $R_0$ in the summer |
| $\varphi$ | -3.8 | Weeks | Phase shift of the seasonal forcing |

## Notes

### Competing Interest Statement

The authors have declared no competing interest.

### Funding Statement

The authors received no funding for this work.

